# Predictors of unfavourable treatment outcome in patients diagnosed with drug-resistant tuberculosis in the Torres Strait / Papua New Guinea border region

**DOI:** 10.1101/2022.03.22.22272751

**Authors:** J’Belle Foster, Diana Mendez, Ben J. Marais, Dunstan Peniyamina, Emma S. McBryde

## Abstract

Drug-resistant tuberculosis (DR-TB) is an ongoing challenge in the Torres Strait Islands (TSI) / Papua New Guinea (PNG) border region. Treatment success rates have historically been poor for patients diagnosed with DR-TB, leading to increased transmission. This study aimed to identify variables associated with unfavourable outcome in patients diagnosed with DR-TB to inform programmatic improvements.

A retrospective study of all DR-TB cases who presented to Australian health facilities in the Torres Strait between 1 March 2000 and 31 March 2020 was performed. This time period covers four distinct TB programmatic approaches which reflect Australian and Queensland Government decisions on TB management in this remote region. Univariate and multivariate predictors of unfavourable outcome were analysed. Unfavourable outcome was defined as lost to follow up, treatment failure and death. Successful outcome was defined as cure and treatment completion.

In total, 133 patients with resistance to at least one TB drug were identified. The vast majority (123/133; 92%) of DR-TB patients had pulmonary involvement; and of these, 41% (50/123) had both pulmonary and extrapulmonary TB. Unfavourable outcomes were observed in 29% (39/133) of patients. Patients living with human immunodeficiency virus, renal disease or diabetes (4/133; 4/133; 3/133) had an increased frequency of unfavourable outcome (*p* <0.05), but the numbers were small. Among all 133 DR-TB patients, 41% had a low lymphocyte count, which was significantly associated with unfavourable outcome (*p* <0.05). We noted a 50% increase in successful outcomes achieved in the 2016 - 2020 programmatic period, compared to earlier periods (OR 5.3, 95% Confidence Interval [1.3, 20.4]). Being a close contact of a known TB case was associated with improved outcome.

While DR-TB treatment outcomes have improved over time, enhanced surveillance for DR-TB, better cross border collaboration and consistent diagnosis and management of comorbidities and other risk factors should further improve patient care and outcomes.

## Introduction

Drug resistance poses a major threat to global tuberculosis (TB) control, with nearly half a million of the ten million cases in 2019 estimated to be rifampicin resistant (1). Drug-resistant (DR)-TB refers to TB strains that are resistant to any of the first-line drugs used to treat fully-susceptible TB, any of the second-line drugs used to treat DR-TB, or are resistant to a combination of these drugs (2). Inadequate adherence to TB treatment may lead to the development of DR-TB and multidrug-resistant (MDR)-TB; resistant to both isoniazid and rifampicin and substantially more difficult and costly to treat (3). Treatment outcomes are generally worse for DR-TB cases due to the use of less potent drugs, prolonged treatment regimens, difficult adherence and potential severe drug-related adverse effects (4). Globally, the average treatment success rate in patients with MDR-TB in 2018 was 59% (5), although this is highly variable across countries (Ukraine – 18.1% (6); China – 52.2% (7); Ethiopia – 78.6%) (8).

TB control poses an ongoing public health challenge in the Torres Strait / Papua New Guinea (PNG) region. The challenge is compounded by high levels of DR-TB in the Western Province of PNG and in particular, on Daru Island (9, 10) (Fig 1). In 2016, the estimated incidence rate of TB in PNG was 432 / 100,000 population (WHO, 11); 674 / 100,000 in the Western Province (12). By comparison, the incidence rate of TB over the border in Queensland, Australia is 5.5 / 100,000 population (13). In the Torres Strait Islands, 80% of people identify as Torres Strait Islander Indigenous peoples (14). Indigenous Australians are disproportionately affected by TB when compared to non-Indigenous Australians (15). It is important to appreciate that residents from specific islands in the Torres Strait and villages in the Western Province of PNG share an open international border (Fig 1), heightening cross-border TB transmission risk (10).

**Fig 1.**
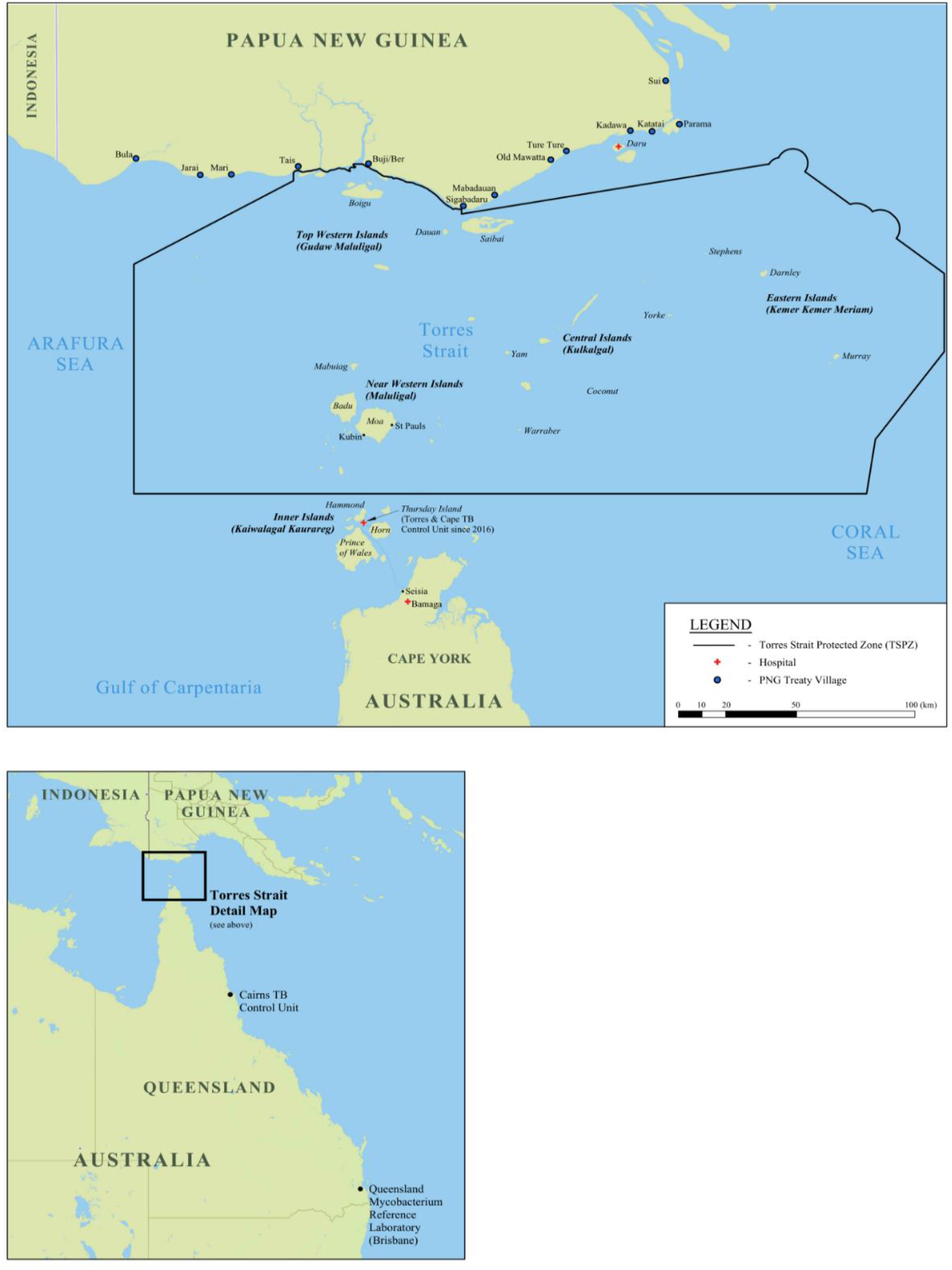
Map of the Torres Strait / Papua New Guinea cross-border region (16)^1^ CC BY 4.0. ^1^International travel without passport or visa is permitted for traditional inhabitants of the Torres Strait Protected Zone and Treaty villages of Papua New Guinea, DOI. 10.6084/m9.figshare.16632823

Management of DR-TB is complex and is often further complicated by comorbidities with other communicable and non-communicable diseases (17). Patients with TB and comorbidities such as renal impairment, diabetes and human immunodeficiency virus (HIV) are more likely to have unfavourable outcomes and are known causes of TB reactivation (18-20). While the rise in DR-TB is a global concern, TB programs must consider specific local risk factors and programmatic gaps to ensure better outcomes for patients. These considerations may present important opportunities for TB programmes to meet the challenging END TB targets, aiming to reduce the incidence of TB by 90% before 2035 (21).

The aim of this study was to evaluate variables associated with unfavourable outcome and the impact of programmatic changes to models of TB care over time with treatment outcomes in DR-TB patients diagnosed in the Torres Strait / PNG border region. It is the intention that evidence derived from this study will be carefully considered at a programmatic level to further improve outcomes for patients diagnosed with DR-TB in this context.

## Methods

### Study Design and Population

We performed a retrospective cohort study of all patients diagnosed with laboratory-confirmed DR-TB between 2000 and 2020 in the Torres Strait Islands, Australia. Pulmonary, extrapulmonary, smear positive and smear negative cases were included, as well as those with mono (isoniazid, rifampicin or streptomycin mono-resistance), poly (resistant to two or more TB drugs) and MDR-TB (resistant to isoniazid and rifampicin). Streptomycin mono-resistance was included, given the high prevalence reported in parts of PNG (22). Ethionamide resistance was analysed in relation to MDR-TB cases, given reports of frequent isoniazid and ethionamide co-resistance amongst MDR-TB strains circulating on Daru Island (23, 24).

Patients were excluded from the study if they were residents of PNG villages external to the Western Province of PNG who did not enter the Australian health system via a health facility in the Torres Strait Protected Zone (Fig 1).

### Models of TB Care – Diagnosis and Treatment

There were four models of TB care provided in the region during the study period, over different periods of time (2000-2005; 2006-2012; 2013-2015; 2016-2020) (25). Supplementary material associated with the models of TB care in this region are available online from https://doi.org/10.6084/m9.figshare.16834648.v1. Across all four models of TB care, the primary mode of diagnosis was via sputum collection. Appropriate specimens for the diagnosis of extrapulmonary TB were rarely collected in these remote Australian border clinics. Mycobacterial culture, phenotypic drug susceptibility testing and genotyping were available throughout the study period and were performed on at least one specimen per patient in the Queensland Mycobacterium Reference Laboratory in Brisbane (Fig 1) (24). The major differences in care resulting from these changes were reduction in time to treatment commencement and retention in care (26).

Between 2000 and 2012, some treatment for DR-TB was available for PNG nationals diagnosed with DR-TB in Australian border clinics, however, access to treatment was improved between 2006 and 2012 with the establishment of frequent outreach TB clinics in the Torres Strait. The change from the 2000-2005 model of TB care to the 2006-2012 model of TB care was in direct response to Australian Government decisions to invest in TB management of PNG patients from two outer Torres Strait Islands, Boigu and Saibai (27). The 2013-2015 model of TB care is reflective of Government decisions to stand-down these outer island clinics and increase support and funding for TB health services on the PNG side of the border (28). As a result, PNG residents diagnosed from 2013 onwards were referred back to the PNG health system for treatment and management. Government decisions to invest in a local Torres Strait-based TB Control Unit with the aim to more rapidly and appropriately respond to presumed TB and confirmed DR-TB cases occurred from 2016, and is ongoing (29). All Australian residents in this study were managed within the Australian public healthcare system.

### Data Collection

The Queensland Health’s Notifiable Conditions System (NoCS) was used as the source for TB notification data. Only cases with laboratory confirmed DR-TB were included in the study. We used Queensland Health’s laboratory software (AUSLAB) as cross-reference. One case was added to the study where their drug-resistant status was identified in AUSLAB, but not registered in NoCS.

Relevant biomarker data (haemoglobin, albumin, lymphocyte levels) were obtained from AUSLAB. Routinely collected biomarker data were analysed, but the fact that it was not consistently available in all patients may have introduced some selection bias.

The management of DR-TB at the Torres Strait / PNG border presents challenges as it involves patients from two countries accessing health systems on both sides of the border in a very remote geographical location. Given the level of interaction between inhabitants, and the increased risk of DR-TB transmission in this cross-border region (9, 10), outcomes for patients with DR-TB from both sides of the border needed to be considered to evaluate the management of DR-TB in this complex context. Hence, the main strength of this study lies in the linkage and analysis of different data sources. Data from the Queensland Department of Health sources (NoCS and AUSLAB) and the local electronic patient information system used in the Torres Strait called ‘Best Practice’ were linked to obtain detailed information for each patient included in this study. This data linkage resulted in a more comprehensive insight into the management of DR-TB in the region.

### Definitions

In this study, we used WHO definitions (31), where unfavourable treatment outcome was defined as death, lost to follow up or treatment failure. Successful treatment outcome was defined as completed treatment or cured. Transferred out, indicated that the patient was referred back to the PNG health care system. In some patients that were transferred out, treatment outcome was unknown.

Comorbidities were defined as HIV infection, diabetes and renal impairment. Patients with comorbidities recorded as renal dysfunction, renal insufficiency, renal disease and renal failure were all defined as having renal impairment.

With reference to haemoglobin levels, anaemia was defined as those with a Z score at least 2 standard deviations (SD) away from the mean, and severe anaemia was defined as those with a Z score that was at least 5SD away from the mean. This resulted in our definition of anaemia fitting with the laboratory definition of below the reference range, for a given laboratory and patient profile. Within laboratory software AUSLAB, the result is documented in an orange-coloured if it is at least 2SD away from the mean (anaemia) and in a red-colour if at least 5SD away from the mean (severe anaemia) (32). Similarly, low albumin and lymphocyte levels were defined as those with a Z score at least 2SD away from the mean.

## Data Analyses

Statistical analyses were performed using IBM SPSS Statistics, version 25 (2019, Armonk, New York, United States). Frequencies and percentages were calculated for descriptive data including age categories, sex, country of birth, visa status, primary health centre (PHC) attended, programmatic diagnosis year group, site of disease, case type, comorbidities, drug resistance, cough and known close contact status. Pearson’s Chi-squared tests were carried out to assess whether age group, sex, country of birth, visa status, and PHC attended were associated with unfavourable treatment outcomes.

Potential categorical predictors (site of disease, HIV infection status and comorbidities, case type, drug resistance and selected biomarkers including haemoglobin, lymphocytes and albumin), were analysed by unfavourable, successful or unknown treatment outcome using Fisher’s exact or likelihood ratios, except for ethionamide resistance and lymphocyte levels where Pearson Chi-Square was used to ascertain if there was a statistically significant association between these factors and treatment outcomes. Likelihood ratios were used to assess the association between unfavourable treatment outcome and more than one categorical variable. The categorical variable ‘diagnostic year group’ was included in the analysis as a clinical covariate to account for programmatic changes in the clinical management of TB over time.

In all univariate and multivariate logistic regression analyses, multiple imputation was applied and pooled results from five imputations were used as 19 DR-TB cases did not have any biomarker results available. Biomarker Z scores were imputed for incomplete variables and were further defined as nominal (unfavourable or other) prior to imputation. To overcome differences in reference range parameters that were automatically applied by Queensland Health’s laboratory software, AUSLAB to biomarker results based on sex and age, Z scores were calculated in Microsoft Excel version 2016.

Univariate analysis of comorbidities, diagnosis year group, contact with a known case, biomarkers, acid-fast bacilli (AFB) positivity and rifampicin-resistance were examined as potential predictors of unfavourable outcome using binary logistic regression. All predictor variables were considered for multivariate regression. The regression method was a forward algorithm with entry criteria of *p* < 0.05 from the univariate analyses followed by a backward algorithm with back entry criteria of *p* > 0.05. The level of significance was set at *p* <0.05 for all analyses.

### Ethics

The Far North Queensland Human Research Ethics Committee (HREC) (HREC/17/QCH/74-1157) granted a waiver of consent and use of anonymized data, and approved the study, as did the Chair of James Cook University HREC, (H7380). Further approval was obtained to access case notification data via a Public Health Act application (QCH/36155 – 1157).

## Results

In total, we identified 133 DR-TB patients during the study period. There were 22 deaths and one treatment failure, 16 were lost to follow up and 51 were transferred out. Of the 51 that were transferred out, 41 had no recorded outcome. As shown in Table 1, Boigu Island PHC received fewer patients than Saibai Island PHC but had marginally greater treatment success (49% vs 38%). The median age was 28 years, and patients in the 15-29 years age group were both the largest group and disproportionately affected by unfavourable treatment outcomes. Of six adults aged ≥60 years, 83% (*n* = 5) had an unfavourable treatment outcome. Being a PNG national rather than an Australian Torres Strait Islander was highly predictive of unfavourable treatment outcome.

**Table 1.** Demographic characteristics and treatment outcome of all patients diagnosed with drug-resistant tuberculosis in the Torres Strait Islands between 2000 and 2020.

| Demographic characteristic | Treatment Outcome<br>N = 133 (%)<sup>†</sup> |  |  |  |  |  |  | p-value |
| --- | --- | --- | --- | --- | --- | --- | --- | --- |
|  | Successful |  | Unfavourable |  |  | Transferred Out |  |  |
|  | Cured | Completed | Died | Failed | Lost to follow up |  | Total |  |
| **Age group** |  |  |  |  |  |  |  | **0.02** |
| <5 years | 2 (22) | 5 (56) | 0 (0) | 0 (0) | 0 (0) | 2 (22) | 9 |  |
| 5-14 years | 1 (10) | 3 (30) | 1 (10) | 0 (0) | 2 (20) | 3 (30) | 10 |  |
| 15-44 years | 9 (9) | 30 (32) | 15 (16) | 0 (0) | 14 (15) | 27 (28) | 95 |  |
| 45-59 years | 0 (0) | 2 (15) | 2 (15) | 0 (0) | 0 (0) | 9 (70) | 13 |  |
| $\geq 60$ years | 1 (17) | 0 (0) | 4 (67) | 1 (17) | 0 (0) | 0 (0) | 6 | |
| **Sex** |  |  |  |  |  |  |  | **0.7** |
| Female | 9 (12) | 22 (30) | 14 (19) | 1 (1) | 8 (11) | 21 (28) | 75 |  |
| Male | 4 (7) | 18 (31) | 8 (14) | 0 (0) | 8 (14) | 20 (35) | 58 |  |
| **Country of Birth** |  |  |  |  |  |  |  | **0.02** |
| Australia | 3 (50) | 2 (33) | 1 (17) | 0 (0) | 0 (0) | 0 (0) | 6 |  |
| Papua New Guinea | 10 (8) | 38 (30) | 21 (17) | 1 (1) | 16 (13) | 41 (32) | 127 |  |
| **Visa Status** |  |  |  |  |  |  |  | **0.002** |
| Australian resident | 3 (33) | 3 (33) | 2 (22) | 1 (11) | 0 (0) | 0 (0) | 9 |  |
| Papua New Guinea Treaty Visitor | 5 (6) | 23 (27) | 15 (17) | 0 (0) | 13 (15) | 30 (35) | 86 |  |
| Papua New Guinea non-Treaty Visitor | 5 (13) | 14 (37) | 5 (13) | 0 (0) | 3 (8) | 11 (29) | 38 |  |
| **Primary Health Centre Attended** |  |  |  |  |  |  |  | 1.0 |
| Saibai Island | 11 (10) | 32 (29) | 19 (17) | 1 (1) | 14 (13) | 33 (30) | 110 |  |
| Boigu Island | 2 (13) | 7 (44) | 2 (13) | 0 (0) | 2 (13) | 3 (19) | 16 |  |
| Murray Island | 0 (0) | 0 (0) | 0 (0) | 0 (0) | 0 (0) | 2 (100) | 2 |  |
| Darnley Island | 0 (0) | 1 (50) | 0 (0) | 0 (0) | 0 (0) | 1 (50) | 2 |  |
| Yorke Island | 0 (0) | 0 (0) | 1 (50) | 0 (0) | 0 (0) | 1 (50) | 2 |  |
| Thursday Island | 0 (0) | 0 (0) | 0 (0) | 0 (0) | 0 (0) | 1 (100) | 1 |  |

<sup>†</sup> Percentages may not equal 100 due to rounding.

Table 2 shows that DR-TB patients diagnosed between 2000 and 2005 were more likely to have unfavourable treatment outcomes (50%; *n* = 7) than those diagnosed between 2016 and 2020 (17%; *n = 2)*. Overall, outcome improved in recent years with a 50% increase in the chance of a successful outcome between 2016 – 2020, when compared to all other programmatic year groups; OR 5.3, 95% Confidence Interval (CI) [1.3, 20.4]. Of 133 DR-TB cases, 67% (*n* = 89) had MDR-TB and of those, 74% (*n* = 66) were new cases and 32% had an unfavourable treatment outcome. Ethionamide resistance was only identified in patients diagnosed with MDR-TB (*p* <.001); 90% (*n* = 80) of MDR-TB cases were found to be ethionamide resistant. Nearly one in five patients had previously received full or partial treatment in Australia or PNG. The composite comorbidity variable comprised of nine patients with DR-TB and HIV infection, diabetes and/or renal impairment. Of four patients with DR-TB/HIV coinfection, and three patients with DR-TB/diabetes comorbidity, none had a successful outcome. Overall, 78% (*n* = 7) of patients with DR-TB and at least one comorbidity had an unfavourable treatment outcome.

**Table 2.**
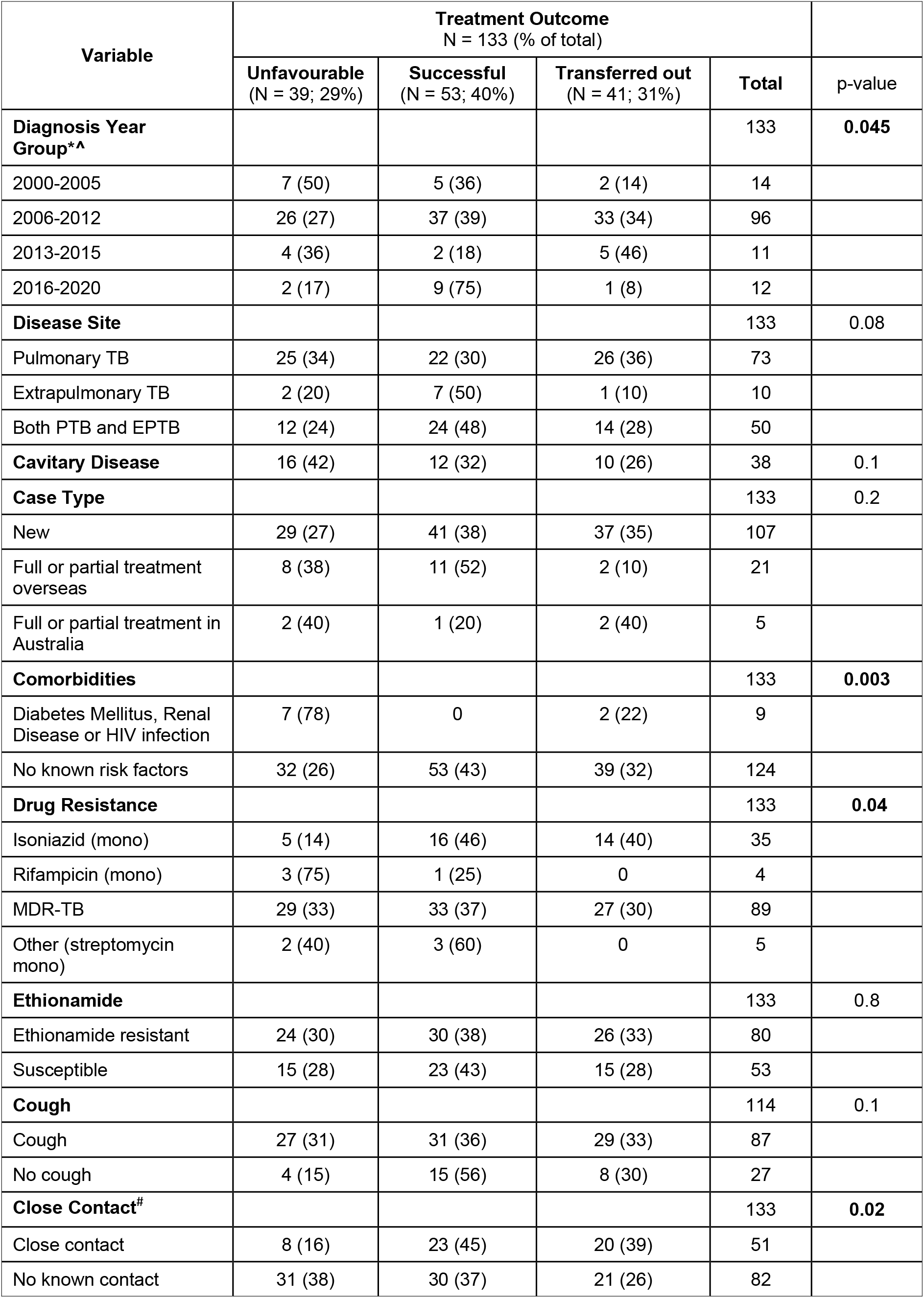
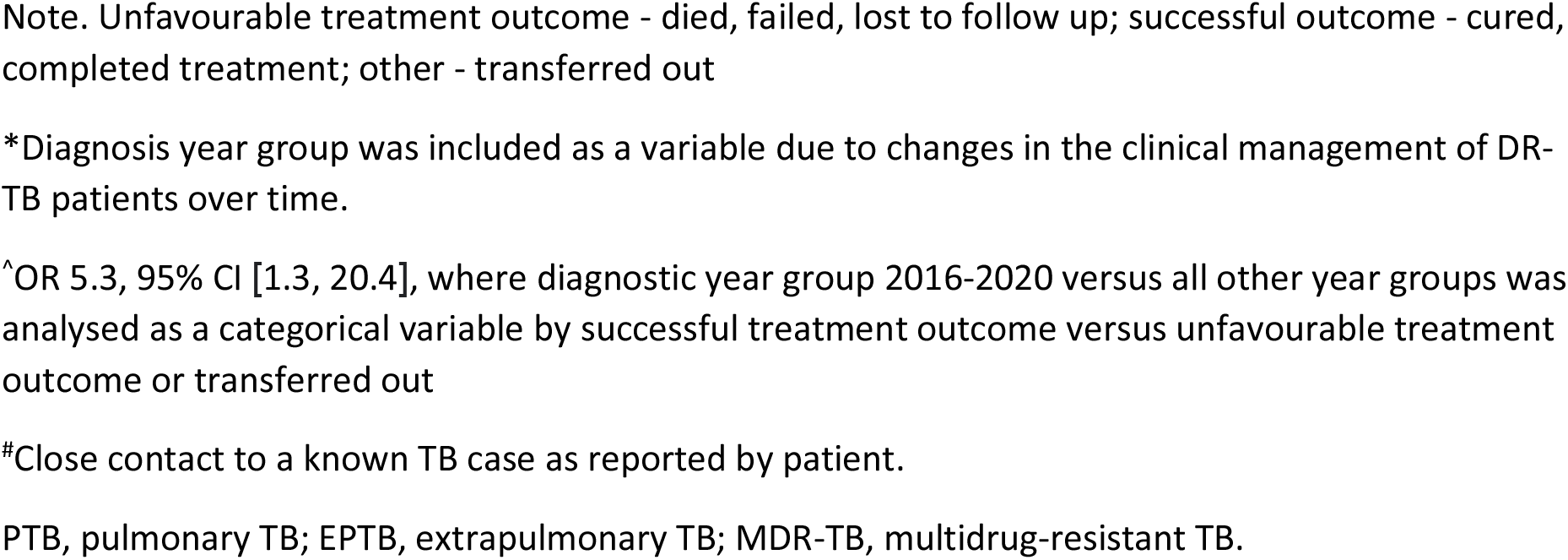
Clinical variables of patients diagnosed with drug-resistant tuberculosis in the Torres Strait between 2000 and 2020, and their association with successful, unfavourable and other TB treatment outcome.

| Variable | Treatment Outcome<br>N = 133 (% of total) |  |  |  | p-value |
| --- | --- | --- | --- | --- | --- |
|  | Unfavourable<br>(N = 39; 29%) | Successful<br>(N = 53; 40%) | Transferred out<br>(N = 41; 31%) | Total |  |
| **Diagnosis Year Group\*^** |  |  |  | 133 | **0.045** |
| 2000-2005 | 7 (50) | 5 (36) | 2 (14) | 14 |  |
| 2006-2012 | 26 (27) | 37 (39) | 33 (34) | 96 |  |
| 2013-2015 | 4 (36) | 2 (18) | 5 (46) | 11 |  |
| 2016-2020 | 2 (17) | 9 (75) | 1 (8) | 12 |  |
| **Disease Site** |  |  |  | 133 | 0.08 |
| Pulmonary TB | 25 (34) | 22 (30) | 26 (36) | 73 |  |
| Extrapulmonary TB | 2 (20) | 7 (50) | 1 (10) | 10 |  |
| Both PTB and EPTB | 12 (24) | 24 (48) | 14 (28) | 50 |  |
| **Cavitary Disease** | 16 (42) | 12 (32) | 10 (26) | 38 | 0.1 |
| **Case Type** |  |  |  | 133 | 0.2 |
| New | 29 (27) | 41 (38) | 37 (35) | 107 |  |
| Full or partial treatment overseas | 8 (38) | 11 (52) | 2 (10) | 21 |  |
| Full or partial treatment in Australia | 2 (40) | 1 (20) | 2 (40) | 5 |  |
| **Comorbidities** |  |  |  | 133 | **0.003** |
| Diabetes Mellitus, Renal Disease or HIV infection | 7 (78) | 0 | 2 (22) | 9 |  |
| No known risk factors | 32 (26) | 53 (43) | 39 (32) | 124 |  |
| **Drug Resistance** |  |  |  | 133 | **0.04** |
| Isoniazid (mono) | 5 (14) | 16 (46) | 14 (40) | 35 |  |
| Rifampicin (mono) | 3 (75) | 1 (25) | 0 | 4 |  |
| MDR-TB | 29 (33) | 33 (37) | 27 (30) | 89 |  |
| Other (streptomycin mono) | 2 (40) | 3 (60) | 0 | 5 |  |
| **Ethionamide** |  |  |  | 133 | 0.8 |
| Ethionamide resistant | 24 (30) | 30 (38) | 26 (33) | 80 |  |
| Susceptible | 15 (28) | 23 (43) | 15 (28) | 53 |  |
| **Cough** |  |  |  | 114 | 0.1 |
| Cough | 27 (31) | 31 (36) | 29 (33) | 87 |  |
| No cough | 4 (15) | 15 (56) | 8 (30) | 27 |  |
| **Close Contact<sup>#</sup>** |  |  |  | 133 | **0.02** |
| Close contact | 8 (16) | 23 (45) | 20 (39) | 51 |  |
| No known contact | 31 (38) | 30 (37) | 21 (26) | 82 |  |

Note. Unfavourable treatment outcome - died, failed, lost to follow up; successful outcome - cured, completed treatment; other - transferred out
\*Diagnosis year group was included as a variable due to changes in the clinical management of DR-TB patients over time.
<sup>^</sup>OR 5.3, 95% CI [1.3, 20.4], where diagnostic year group 2016-2020 versus all other year groups was analysed as a categorical variable by successful treatment outcome versus unfavourable treatment outcome or transferred out
<sup>#</sup>Close contact to a known TB case as reported by patient.
PTB, pulmonary TB; EPTB, extrapulmonary TB; MDR-TB, multidrug-resistant TB.

Table 3 shows that 86% (*n* = 92) of patients with haemoglobin recorded had anaemia, and of those, 38% (*n* = 35) had severe anaemia. Low albumin was detected in 76% (*n* = 80) of patients, where this was measured. A large percentage of patients with low lymphocyte levels, (44%; *n* = 43) had an unfavourable treatment outcome.

**Table 3.** Blood test abnormalities in patients diagnosed with drug-resistant tuberculosis in the Torres Strait between 2000 and 2020, and their association with treatment outcome.

| Variable | Treatment Outcome n (%) |  |  |  |
| --- | --- | --- | --- | --- |
|  | Unfavourable | Other | Total | p-value |
| **Haemoglobin Levels\* (50-146L)** |  |  | 107 | 0.5 |
| No anaemia | 3 (20) | 12 (80) | 15 |  |
| Mild anaemia | 20 (35) | 37 (65) | 57 |  |
| Severe anaemia | 10 (29) | 25 (71) | 35 |  |
| **Lymphocyte count# (0.04-9.80L)** |  |  | 106 | **0.01** |
| Normal | 13 (21) | 50 (79) | 63 |  |
| Low | 19 (44) | 24 (56) | 43 |  |
| **Albumin# (<15-48L)** |  |  | 105 | 0.07 |
| Other (normal or high) | 4 (16) | 21 (84) | 25 |  |
| Low | 28 (35) | 52 (65) | 80 |  |

\*Mild anaemia, at least 2 standard deviations (SD) away from the mean; severe anaemia, at least 5SD away from the mean; #Low lymphocyte and albumin, at least 2SD away from the mean.

In Table 4, patients with comorbidities, low lymphocyte levels and AFB positivity were significantly more likely to have unfavourable treatment outcomes. Being a close contact of a known TB case was a protective factor and reduced the odds of an unfavourable treatment outcome occurring (*p* .008; OR .31). Although the p value for anaemia in Table 3 was > .1, this variable was included in the univariate analysis as other studies have demonstrated an association between anaemia and unfavourable treatment outcomes (33), however in this study, anaemia was not a significant predictor of unfavourable treatment outcomes. In the adjusted multivariate model, comorbidities, being a close contact of a known TB case and low lymphocyte levels retained significance (*p* <0.05).

**Table 4.**
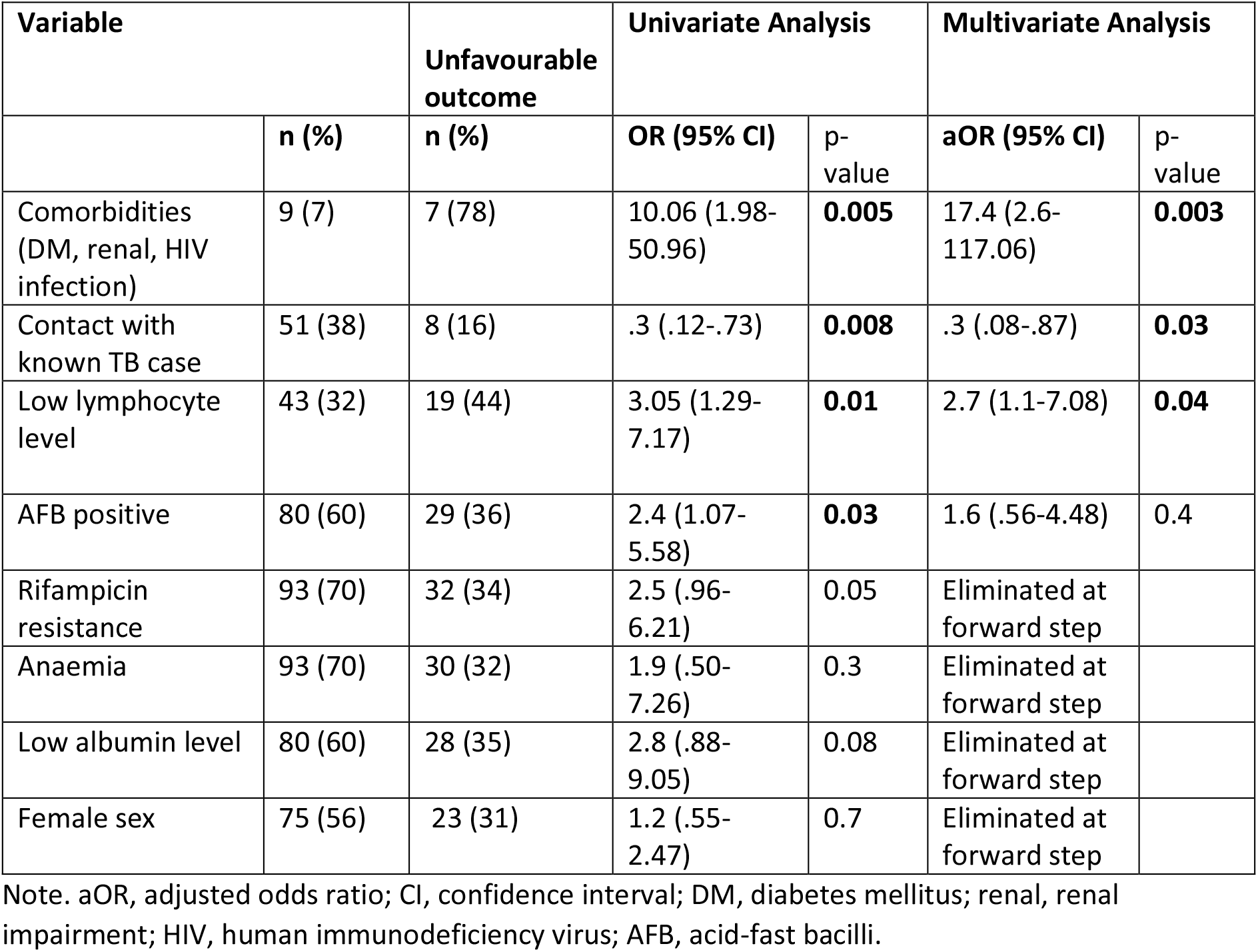
Variables associated with unfavourable treatment outcome among drug-resistant tuberculosis cases diagnosed in the Torres Strait / Papua New Guinea border region between 2000 and 2020 (N = 133)

## Discussion

Few reports focus on the specific challenges and unique health care access issues experienced by DR-TB patients in remote settings like the Torres Strait / PNG border region. This study identified variables associated with unfavourable treatment outcomes in patients diagnosed with DR-TB and examined the impact of four different models of TB care over two decades. We documented a significant improvement in successful treatment outcomes once a local decentralised TB control unit was established in 2016. Patients diagnosed before 2013 had the worst treatment outcomes, which is reflective of a time when accessibility to mycobacterial culture and drug susceptibility testing was not routinely available on Daru Island in PNG, and where access to second-line TB drugs was only available for Western Province PNG nationals via Australian TB clinics (34). For patients diagnosed between 2013 and 2015 in this study, a higher proportion had an unknown outcome. This is consistent with the changeover from the 2006 to 2012 TB management model of increased surveillance and detection at border clinics, to a handover period whereby PNG patients diagnosed at Australian border health facilities were referred back to the PNG health system for ongoing management and care.

From 2016 onwards, TB clinicians were based in the Torres Strait and therefore able to rapidly respond to cases diagnosed, associated contact tracing efforts and monitoring of treatment compliance and outcomes for patients. Higher success rates for DR-TB patients have been reported in countries where patients have access to developed health infrastructure and where skilled clinicians are positioned to support DR-TB patients (35). From 2016, local nurses and Indigenous Health Workers undertook Directly Observed Therapy for TB patients residing in the Torres Strait. Collegial relationships between TB programs in the Torres Strait and Daru Island were also strengthened through this period with the joint development of procedural documents and processes (36) which enabled each TB program to define data requirements, streamline the exchange of shared patient information, and enhance surveillance capability. These strengthened relationships may help to explain the reduction in unknown treatment outcomes in this study from 2016. These initiatives are consistent with the Australian National Tuberculosis Advisory Committee (NTAC) recommendations to engage in regional and bilateral collaborations in order to improve TB services in cross-border areas (37).

It is widely accepted that household contacts of DR-TB patients are at greater risk of exposure, infection and disease progression when compared with other types of contacts (38). In a study of MDR-TB index cases in Pakistan, MDR-TB diagnoses were reported in 17.4% of close contacts (38). Unlike isoniazid-resistant TB which is more amenable to treatment, patients with MDR-TB may be more likely to remain infectious longer than patients with drug susceptible TB (39), thus increasing the likelihood of transmission to close contacts. Despite the increased transmission risk, close contacts had more favourable outcomes in this study. A possible explanation is that these close contact patients were actively screened, or linked in with healthcare services earlier because of raised awareness, leading to earlier diagnosis and potentially improved treatment adherence. Better outcomes for close contacts after 2014 may also be attributed to improved health literacy and symptom recognition as a result of mass community education offered in the region from 2014 (40).

This retrospective study has a number of limitations. Details of the drug regimens used for individual patients in this study were not available for analysis, however twice and thrice weekly dosing of second-line drugs was available for some MDR-TB patients treated between 2006 and 2015. From 2016, clofazimine and linezolid were included in most MDR-TB regimens in the region per WHO recommendations (41), and bedaquiline was first used in the region in 2018. The study did not identify the type of close contact (i.e. household contact), nor ascertain the type of TB that each close contact was exposed to. The recent upgrade of the Mabadauan health centre in PNG will aid the PNG health system’s capacity to manage close contacts in border communities. It will, however, be important that expanded diagnostic capabilities at Mabadauan, are matched with capacity to retain patients in the TB care pathway (35). Since 2016, the Torres and Cape TB Control Unit has collected and shared TB contact tracing data with Daru General Hospital, related to all PNG residents diagnosed with TB in the Torres Strait. It is anticipated that increased capacity of local health services available to residents of PNG living adjacent to the Torres Strait, may lead to effective TB contact management, further improve treatment outcomes for PNG patients diagnosed with DR-TB as well as support TB patients with other comorbidities.

Patients with pre-existing comorbidities in this study were significantly more likely to have unfavourable treatment outcomes, which is consistent with earlier findings (6, 42, 43). In our study, no patients with a serious comorbidity were cured or completed treatment. HIV is a major risk factor for TB disease development and associated with unfavourable treatment outcome in the absence of successful HIV care, which is a major concern in the study setting. However, HIV is unlikely to be a major contributing factor in community TB transmission dynamics as currently less than 1% of the population is HIV infected (44). It is possible that ‘stand-alone’ vertical disease programs for TB control, diabetes management and sexual health, may contribute to poor linkage of care and unfavourable treatment outcomes for DR-TB patients with a comorbidity (45). It would be beneficial for TB programs to establish strong linkages with both diabetes educators and sexual health providers to provide enhanced screening, monitoring and management support for patients with these comorbidities (46). As there were no protocols in places to consistently screen TB patients for diabetes or renal impairment during this study period, it is possible that comorbidities have been under-reported in this study.

In renal patients, delayed TB diagnosis is a possible reason for unfavourable outcome (47). Aboriginal and Torres Strait Islander peoples develop chronic kidney disease three times as often as non-Indigenous Australians (48) and in Torres Strait Islander adults, a recent survey found that nearly one in five people showed signs and symptoms of chronic kidney disease (49). Renal impairment is more likely to be documented in residents of the Torres Strait Islands due to their relative ease of access to ongoing health services in the region. By contrast, an absence of renal dialysis in the Treaty villages suggests there would be limited survival of PNG patients with kidney disease. As uremic symptoms like fever and weight loss are non-specific symptoms of both TB and renal disease, renal physicians would be well placed to consider TB as a differential diagnosis and monitor patients with insidious onset of these symptoms (47). Early detection of TB in renal patients may be key to improved outcomes (47). Efforts to increase collaboration between renal and TB units in the Torres Strait may help reduce diagnostic delay and better support shared patients. Many studies have reported high rates of mortality in TB patients with comorbidities (47, 50-52) and in patients with low levels of haemoglobin, albumin and lymphocytes (53).

Although not consistently measured, low lymphocyte levels were statistically significantly associated with unfavourable treatment outcomes in DR-TB patients, and also among HIV uninfected patients, which is consistent with findings in other studies (54). In patients who were previously lost to follow up or with past treatment failure, overall health decline evidenced by low lymphocyte levels can be a marker for disease severity and complexity (55). When compared to drug susceptible TB, lower lymphocyte counts have been reported in MDR-TB patients (56). Low lymphocyte counts, as well as haemoglobin and albumin levels, are all indicative of general poor health at the time of treatment initiation.

Low haemoglobin and albumin levels have been identified as strong predictors of TB mortality (57) and in China and Israel, an association between hypoalbuminaemia and poor prognosis in DR-TB patients has been reported (7, 58). A study conducted in Ethiopia reported that MDR-TB patients with low haemoglobin levels (anaemia) were more than twice as likely to have unfavourable treatment outcomes when compared to patients without anaemia (42). Low haemoglobin has also been associated with unfavourable treatment outcomes in non-MDR TB patients, specifically with delayed sputum conversion at two months (59). It is possible that the anaemia observed in DR-TB patients is due to nutritional iron deficiency and not chronic disease (33), however this study did not explore levels of iron, ferritin, hepcidin and transferrin in TB patients or assess haemoglobin levels in the general population.

Several limitations have already been described above. Another limitation of this study is that only people with laboratory-confirmed DR-TB isolates were included, excluding those with drug susceptible TB or unconfirmed DR-TB. This therefore represents a very select group and limits our ability to comment on general trends and outcomes. We were however able to compare those with and without rifampicin resistance, finding a trend to worse outcome in those with rifampicin resistance.

Apart from selection bias, the relatively small number of patients also affected the power of our statistical analyses. Future studies could incorporate drug-susceptible patients as well, which would provide a more generalizable comparator. We were unable to ascertain whether the cause of death for all patients was TB, however, patient death was documented during the time period for the course of TB treatment for all patients. As this was an observational study of care delivered over time, we acknowledge that there may be confounding factors, linked to changes in clinical care and the availability of new diagnostic tools and treatment options.

## Conclusion

Although having a locally-based TB program in the Torres Strait has resulted in better treatment outcomes for DR-TB patients, more interventions to improve treatment outcomes and refine the focus on vulnerable sub-populations are needed. Most patients with comorbidities in this study had an unfavourable treatment outcome and poor health indicators suggesting late presentation or underlying poor management of their comorbidity. These findings have important implications for TB programs on both sides of this international border. Key strategies should include strong cross-border working relationships, as well as better collaboration with diabetes, renal and sexual health care providers to identify and manage TB in patients. There should be an ongoing commitment to enhance HIV care and to improve the care of DR-TB patients at-risk of unfavourable treatment outcomes.

## Data Availability

Data cannot be shared publicly due to confidentiality clauses surrounding the use of notifiable disease-based data. Data are available from Queensland Health and require approval from Far North Queensland Human Research Ethics Committee and Public Health Act authorization for researchers who meet the criteria for access to confidential data.

https://doi.org/10.6084/m9.figshare.16632823

